# Monocytes are the main source of STING-mediated IFN-α production

**DOI:** 10.1101/2022.03.11.22272208

**Authors:** Nicolas Congy-Jolivet, Claire Cenac, Jérôme Dellacasagrande, Bénédicte Puissant-Lubrano, Pol André Apoil, Kevin Guedj, Flora Abbas, Sophie Laffont, Sandrine Sourdet, Sophie Guyonnet, Fati Nourhashemi, Jean-Charles Guéry, Antoine Blancher

**Affiliations:** Laboratoire d’Immunologie, CHU Purpan, Toulouse, France; Institut Toulousain des Maladies Infectieuses et Inflammatoires (INFINITY), Université de Toulouse, INSERM, CNRS, Université Paul Sabatier, Toulouse, France; Blood Assay Solutions, 31670 Labège, France; Gérontopôle de Toulouse, Département de Médecine Interne et Gérontologie Clinique, CHU de Toulouse, Toulouse, France; Maintain Aging Research team, CERPOP, Université de Toulouse, INSERM, Université Paul Sabatier, Toulouse, France

**Author notes:** Corresponding authors: Antoine Blancher, MD PhD, Laboratoire d’immunologie, Hôpital Purpan, Institut Fédératif de Biologie, 330 Avenue de Grande Bretagne, TSA 40031, 31059 Toulouse Cedex 9, France or Jean-Charles Guéry, PhD, Institut Toulousain des Maladies Infectieuses et Inflammatoires (INFINITY), INSERM UMR1291, Centre Hospitalier Universitaire Purpan, Place du Dr Baylac, 31024 Toulouse Cedex 3, France. These authors contributed equally to this work (Nicolas Congy-Jolivet, Claire Cenac).

**Keywords:** age, sex, nucleic acid sensing, type I interferons, TLR7, TLR9, plasmacytoid dendritic cells, STING, monocytes

## Abstract

**Background:** Type I interferon (IFN-I) production by plasmacytoid dendritic cells (pDCs) occurs during viral infection, in response to Toll-like receptor 7 (TLR7) stimulation and is more vigorous in females than in males. Whether this sex bias persists in ageing people is currently unknown. In this study, we investigated the effect of sex and aging on IFN-α production induced by PRR agonist ligands.

**Methods:** In a large cohort of individuals from 19 to 97 years old, we measured the production of IFN-α and inflammatory cytokines in whole-blood upon stimulation with either R-848, ODN M362 CpG-C, or cGAMP, which activate the TLR7/8, TLR9 or STING pathways, respectively. We further characterized the cellular sources of IFN-α.

**Findings:** We observed a female predominance in IFN-α production by pDCs in response to TLR7 or TLR9 ligands. The higher TLR7-driven IFN-α production in females was robustly maintained across ages, including the elderly. The sex-bias in TLR9-driven interferon production was lost after age 60, which correlated with the decline in circulating pDCs. By contrast, STING-driven IFN-α production was similar in both sexes, preserved with aging, and correlated with circulating monocyte numbers. Indeed, monocytes were the primary cellular source of IFN-α in response to cGAMP.

**Interpretation:** We show that the sex bias in the TLR7-induced IFN-I production is strongly maintained through ages, and identify monocytes as the main source of IFN-I production via STING pathway.

**Funding:** This work was supported by grants from Région Occitanie/Pyrénées-Méditerranée (#12052910, Inspire Program #1901175), University Paul Sabatier, and the European Regional Development Fund (MP0022856).

**Research in context:** *Evidence before this study:* Type I interferon (IFN-I) production by plasmacytoid dendritic cells (pDCs) occurs during infection with viruses, including SARS-CoV-2, in response to Toll-like receptor 7 (TLR7) stimulation. Early type I IFN production by pDCs in the respiratory tract through TLR7 activation is protective in severe COVID-19. The capacity of female pDCs to produce higher levels of interferon α (IFN-α) in response to TLR7 ligands, compared to those of males, is one immune characteristic that robustly distinguishes the two sexes in middle-aged adults. It is currently unknown whether the superior ability of female pDCs to produce IFN-I upon TLR7 stimulation is maintained with age. In this study, we investigated the impact of sex and aging on the release of innate cytokines (IFN-α, IFN-γ, IL-1β, IL-6, IL-8, TNF-α, MCP1) in a whole-blood assay from 310 healthy volunteers (145 males and 165 females) from 19 to 97 years old, upon stimulation with either TLR7-, TLR9-ligands or with cGAMP, the natural product of cGAS which activates STING (Stimulator of IFN Gene) and has been reported to exhibit potent anti-tumor and adjuvant effects through induction of IFN-I by ill-defined cellular sources.

*Added value of this study:* We observed that IFN-α responses to TLR7 and TLR9 ligands were the only whole blood assay variables exhibiting sex differences among all 21 variables investigated (seven analytes analyzed after stimulation by three different ligands). Our results show that the accrued female response in the TLR7-induced IFN-α production was robustly maintained over ages, including elderly subjects >80. In contrast, STING-induced IFN-I production was similar in both sexes and was maintained with aging possibly as a consequence of the age-related increase in circulating monocyte numbers. Indeed, we demonstrate for the first time that monocytes represent the main cellular source of IFN-I upon cGAMP stimulation of PBMCs.

*Implications of all the available evidence:* This study demonstrates that the heightened TLR7 ligand-induced IFN-α secretion by blood pDCs from females, compared to those from males, is maintained in elderly women, supporting the hypothesis that this pathway could contribute to enhanced protection against virus infections such as SARS-CoV-2 in females. This work also shows that cGAMP can promote IFN-I production by targeting monocytes, which numbers increase with aging, suggesting that STING ligands may be useful for vaccine design in the elderly in both sexes.

## Introduction

Biological sex is a modifier of the immune responses against a number of viruses, including influenza [1, 2], HIV-1 [3, 4] and severe acute respiratory syndrome coronavirus 2 (SARS-CoV-2) [5, 6]. In general, women tend to develop stronger innate and adaptive immunity to viral infections, but are at greater risk than men of developing most autoimmune diseases [1, 7]. Emerging evidence support the notion that sex hormones and sex-chromosome loci regulate biological pathways common to autoimmune and infectious diseases [7-10]. This is particularly clear for Toll Like Receptor (TLR)-7, a single-stranded RNA (ssRNA) receptor encoded by an X-linked gene. The response of innate immune cells and B cells initiated by the TLR7-mediated sensing of ssRNA is an essential line of defense against exogenous RNA viruses [11-14] and endogenous retroviruses [15, 16]. Moreover, TLR7 can also respond to endogenous ssRNA containing ligands, potentially leading to autoimmunity or inflammation if not properly controlled [17].

The capacity of female plasmacytoid dendritic cells (pDCs) to produce higher levels of interferon α (IFN-α) in response to TLR7 ligands, compared to those of males, is one immune characteristic that robustly distinguishes the two sexes in middle-aged adult humans [18-21] as well as prepubescent young people [22]. Among cells of innate immune system, pDCs are the major producers of type I IFN in the blood and constitutively express TLR-7 and -9, that sense ssRNA and DNA with a CpG motif, respectively [23]. These endosomal TLRs are responsible for the detection of viral and endogenous nucleic acids and trigger the release of type 1 IFN, including thirteen IFN-α subtypes and IFN-ß [24, 25]. All type I interferon family members activate their common receptor promoting the expression of IFN-stimulated genes (ISGs) in immune and non-immune cells to fight viral infections [23, 24]. Sex differences in TLR7-driven interferogenesis have been suggested to contribute to the observed sex bias in HIV-1 disease progression [4], the enhanced female resistance to COVID-19 [5] and the increased female susceptibility to systemic lupus erythematosus (SLE) [20, 26]. Whether the superior capacity of TLR-stimulated pDCs from female to produce IFN-I is maintained with age is currently unknown.

In this study, we investigated the effect of sex and aging on IFN-α production induced by PRR agonists in a relatively large cohort (n=312) of healthy donors equilibrated for both sexes from 19 to 97 years old. We chose to use a whole blood stimulation assay as such system showed greater reproducibility for monitoring innate immune cell activation [27, 28]. We measured the production of multiple cytokines, including IFN-α, in response to TLR7/8 and TLR9 ligands, which have been validated to induce the production of IFN-I by blood pDCs [29-32]. As an alternative signaling pathway to induce IFN-I, we investigated in parallel the impact of sex and aging on the production of IFN-α induced by cGAMP, the natural product of cGAS which activates the STING (Stimulator of IFN Gene) pathway [33]. Lastly, we explored the main cellular source responsible for the production of IFN-α in human PBMCs stimulated with endosomal TLR7/8/9 ligands or the STING natural ligand cGAMP.

## Methods

### Individuals, blood collection and study approval

Peripheral blood venous samples were collected from 167 healthy donors (83 males and 84 females) from the French Blood Establishment (EFS: Etablissement Français du Sang), or from 143 subjects (62 males and 81 females over 60 years old,) seen in consultation at the “ Gérontopôle” of the Toulouse University Hospital for an overall clinical evaluation aimed to propose them a personalized age-related risk prevention plan. Elderly patients with impaired higher cognitive functions were excluded from the protocol. All participants gave their written informed consent and filled a questionnaire to check the absence of exclusion criteria (ongoing or chronic infection; antibiotherapy treatment for less than 2 months; chronic treatment by corticosteroids, non-steroidal anti-inflammatory drugs, immunosuppressant, immunomodulators, chemotherapy; allergy; vaccination less than three months before the visit; diabetes; acquired or constitutive immunodeficiency; chronic inflammatory or autoimmune disease). For each participant, blood was collected in one EDTA anticoagulated tube and one heparinized tube. Blood samples were processed at the laboratory within 4 hours after bleeding.

### Flow cytometry analysis

Flow cytometry analyses were performed on whole blood within 4 h after blood drawing. Absolute numbers of DC and monocytes were obtained using True Count tubes. 200 µL of whole blood were stained with following antibodies all from BD Biosciences: CD14-APC-H7 (Cat# 641394, RRID:AB_1645725), CD16-V500 (Cat# 561394, RRID:AB_10611857), CD64-V450 (Cat# 561202, RRID:AB_10564066), CD163-FITC (Cat# 563697, RRID:AB_2738379), CD11c-PE (Cat# 555392, RRID:AB_395793), CD123-PE-Cy7 (Cat# 560826, RRID:AB_10563898) and HLA-DR-APC (Cat# 559866, RRID:AB_398674). A mix of PerCP-Cy5.5 conjugated antibodies specific for CD3 (Cat# 560835, RRID:AB_2033956) CD19 (Cat# 561295, RRID:AB_10644017) CD20 (Cat# 558021, RRID:AB_396990) CD56 (Cat# 560842, RRID:AB_2033964) and CD66a 5 (Cat# 562254, RRID:AB_11154419) was used to exclude Lin^+^ cells (granulocytes, lymphocytes T, B, and NK) from the analysis. Monocytes were defined as Lin^-^CD14^+^ cells; DCs were identified as Lin^-^CD14^-^HLA-DR^+^CD123^-^CD11c^+^; pDCs were identified as Lin^-^CD14^-^HLA-DR^+^CD11c^-^CD123^+^. After lysis of erythrocytes with the BD FACS Lysis solution (BD Biosciences Cat# 349202, RRID:AB_2868862) samples were acquired on a FACSCanto IITM flow cytometer, RRID:SCR_018056 (BD Biosciences) and FlowJo software (Tree Star) was used for data analysis. Details concerning the determination of absolute numbers /mm^3^ of cell and cell subsets of interest are described by Puissant Lubrano et al [34].

### Whole blood functional tests

Stock solutions of pattern recognition receptor (PRR) ligands (Resiquimod (R848) (Cat# tlrl-r848), ODN M362 (a class C CpG ODN, (Cat# tlrl-m362-5) and Cyclic [G(2’,5’)pA(3’,5’)p] cGAMP, (Cat# tlrl-nacga23-1) all purchased from InvivoGen, Toulouse, France) were prepared in RPMI. Fifteen µl of each 10X PRR ligand solution were distributed in 200 µl PCR tubes and stored at-20°C. On tube containing 15 µL of RPMI (Invitrogen, Cat# 31870074) without PRR ligand was used as negative control. The final concentration of the PRR ligands was R848 (1µM), ODN M362 (5 µM) and cGAMP (20 µg/ml). Less than four hours after bleeding, heparinized blood was diluted (1/2) with RPMI before being distributed (135 µl) in the tubes containing the various stimulus. After 24 hours of incubation at 37°C, the supernatants (i.e. plasma diluted with RPMI) were harvested and stored at -80°C until cytokine assays.

### Cytokine dosage

The concentrations of six cytokines (interferon gamma (Cat #HIFNGPET), IL-6 (Cat# 62HIL06PET), IL-8 (Cat# 62HIL08PET), MCP1 (Cat# 62HCCL2PEG), IL-1β (Cat# 62HIL1BPET), TNF-α (Cat# 62HTNFAPET) were quantified in the supernatants using the HTRF^®^ technique (Cisbio Bioassays, France). The IFN-α assay was carried out by ELISA (Mabtech, Sweden, Cat# 3425-1H-6) following the manufacturer instructions.

### CMV serostatus

The anti-CMV IgG and IgM were assessed by chemiluminescence (Architect, Abbott :CMV IgM Cat# 6-C16; CMV IgG Cat# 6C15) with positive thresholds of 6.0 arbitrary units (AU)/ml and 1.0 AU/ml for IgG and IgM, respectively. One hundred ninety-two individuals out of 310 were positive for anti-CMV IgG. The range of IgG anti-CMV was 23.6 to >250 AU/ml in CMV+ individuals.

### Genomic DNA extraction and SNP genotyping

Genomic DNA was extracted from cryopreserved or freshly isolated PBMCs using a NucleoSpin Tissue kit (Macherey-Nagel, Cat# 740952.250) and genotyped for *TLR7* SNPs rs179008, rs3853839, and for the *CXOrf21*/*TASL* SNP rs887369 using KASP allele-specific PCR assays (LGC Genomics, Cat# KBS-2100-100, Cat# KBS-1016-002) run on a LightCycler 480 instrument II (Roche, RRID:SCR_020502) as previously described [21].

### Monocyte or pDC depletion assay

PBMC were separated from whole blood by Pancoll (PAN-Biotech, Cat# P04-60500) density gradient centrifugation. Sorting was performed using autoMACS Pro System (Miltenyi Biotech) according to manufacturer’s instructions. PBMC were stained with CD14-PE antibody (Immunotools, Cat# 21620144, clone 18D11) followed by incubation with anti-PE microbeads (Miltenyi Cat# 130-048-801). The depleted fraction was collected and resuspended in R10 medium – i.e., RPMI 1640 (Sigma-Aldrich) containing 10 % heat inactivated FBS (Sigma-Aldrich), 100 U/ml penicillin, 100 U/ml streptomycin and 2 mM L-glutamine (Life Technologies), 1 mM sodium pyruvate, 100 U/ml non-essential amino-acids, 50 μM 2-mercaptoethanol (Invitrogen). 5 × 10^5^ cells/well were seeded in 96-well plates and stimulated with 20 µg/ml cGAMP or 1 µg/ml R848 (Invivogen France). For pDC depletion, PBMCs were stained with PE conjugated anti-BDCA4 (BioLegend Cat# 354504, RRID:AB_11219194) and anti-CD123 (BD Biosciences Cat# 555644, RRID:AB_396001) antibodies, and then sorted using anti-PE microbeads (Miltenyi) as above. Mock-depleted or pDC-depleted PBMCs were stimulated with R848 or ODN M362. Cell depletion was checked by flow cytometry on LSRII cytometer (BD Biosciences, RRID:SCR_002159). Supernatants were collected at 24 hours and tested for IFNα by ELISA as described above.

### Positive selection of CD14^+^ monocytes

CD14^+^ cells were purified from frozen PBMC using CD14-PE antibody (Immunotools) and EasySep™ Human PE positive selection kit (Stemcell, Cat# 17654) according to manufacturer protocol. Positive fraction containing CD14 cells was resuspended in R10 medium and 3.10^5^ cells were seeded in 96-well plates and stimulated with 20 µg/ml of cGAMP (Invivogen France). IFN-α was measured by ELISA in 24 hours supernatants. Cell purity was checked by flow cytometry, as above.

### Statistical analysis

Statistically significant differences between 2 groups were determined using two-tailed unpaired or paired Student’s *t*-test, Mann-Whitney test or paired Wilcoxon signed-rank test. Multiple comparisons were performed using one-way ANOVA test followed by Sidak’s multiple comparison test or using the non-parametric Kruskall-Wallis test corrected for multiple comparisons as indicated. Values are reported as individual values, and plotted as mean±SEM, or as median and IQ range. Correlation test were performed by means of Pearson test and corrected by using the False Discovery Rate (FDR) as described by Benjamini-Hochberg. Statistical analyses were performed using XLSTAT (RRID:SCR_016299, Addinsoft, Bordeaux, France), and Prism 9 (RRID:SCR_002798, GraphPad Software, La Jolla, CA, USA). All statistical tests were two-tailed and p-values <0.05 were considered to be statistically significant.

### Study approval

Our study is in agreement with applicable French regulations and with the ethical principles of the Declaration of Helsinki. The protocol and the informed consent form were submitted and approved by the local ethic committee (Comité de protection des personnes Sud-Ouest et Outre-mer II, demande DC-2014-2098). We obtained any individual’s free prior informed consent.

## Results

### Study design, cohort description and immune monitoring strategies

We recruited 310 healthy volunteers (165 women and 145 men) whose ages span 19-97 years old (Figure 1a). The study was designed to include similar numbers of males and females for the various age classes and to equilibrate the numbers of older subjects (60+: 63 males, 82 females) compared to young and middle-aged subjects (ages 19-59; 82 males, 83 females). Excepted for the older subjects (ages 80-97: 41 males, 60 females), donors were evenly distributed by age and sex. All individuals were apparently healthy with medical history devoid of known immunodeficiency or immunosuppressive treatment. PBMCs were profiled by flow cytometry to quantify the numbers of white blood cells, including polymorphonuclear neutrophils, lymphocytes, monocytes, conventional DCs and pDCs as previously described [34]. In parallel, we designed an *in vitro* cytokine release assay with human whole blood as described in materials and methods (Figure 1b). We selected three different stimuli known to induce IFN-α production in addition to a large spectrum of inflammatory cytokines (Supplementary Figure S1): R848 a ligand of TLR7 and TLR8, ODN M362 (a CpG-C class ODN) that activates TLR9 and IFN-I production by pDCs [35], and cGAMP which activates the STING pathway. Cytokine concentrations in plasma were measured after 24 h of stimulation an expressed as log of the concentration measured as shown in Supplementary Figure S1. The distribution of log cytokine concentrations fitted a Gaussian distribution. Excepted for IL-1ß and TNF-α production induced by cGAMP, the frequencies of non-responding individuals were similar between men and women (Supplementary Figure S2). For each ligand, we selected a set of analytes giving the highest frequencies of responders between both sexes and ages (Figure 1c-e; Supplementary Figure S1 and S2).

**Figure 1.**
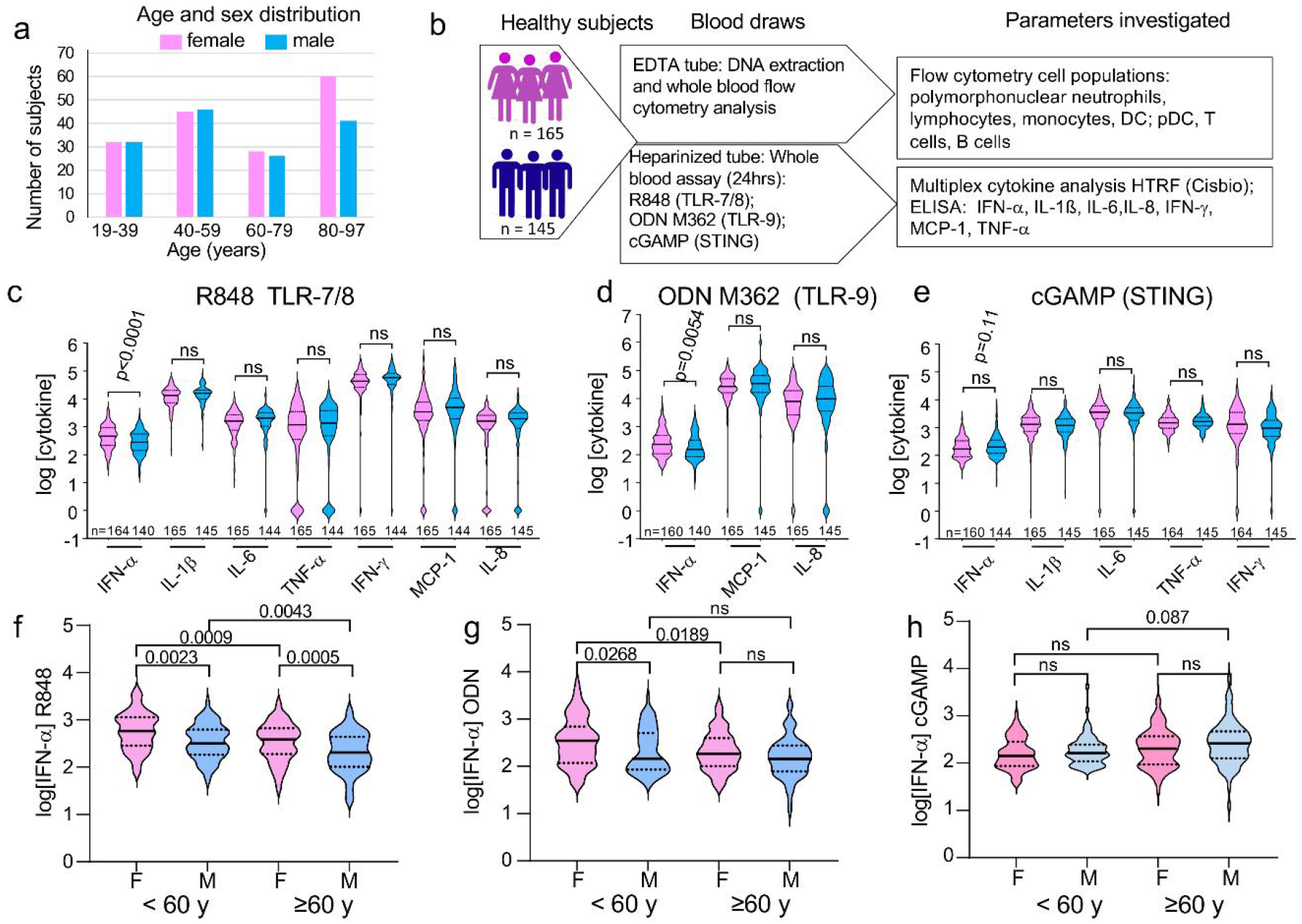
Investigating the sexual dimorphism in the TLR7-, TLR9- and STING-mediated innate cytokine responses with aging. (a) demographic data of the study cohort, age and sex distribution. (b) Schematic summary of study design: Blood samples were obtained from 310 healthy community-dwelling individuals (ages 18-97), samples were separated in two for whole blood stimulation with the indicated PRR ligands, and the other part was used to isolate PBMCs for FACS analysis as indicated. (c, d, e) Levels of cytokine release after whole blood stimulation with the three studied PRR ligands: R848 (c), ODN M362 (d) or cGAMP (e). Concentrations of indicated cytokines were quantified at 24 hours in plasma. The results are given in log_10_ of the concentrations (pg/ml) with background (without ligands) subtracted. (c, d, e, f-h) Sex-differences in IFN-α release in response to R848 (f), ODN M362 (g) and cGAMP (h), before and after the age of 60. Statistical differences between groups were analyzed using the unpaired *Student’s t*-*test (c-d) or* by a one-way ANOVA test followed by Sidak’s multiple comparisons test (f-h).

### Female sex bias in IFN-α production in response to TLR7- or TLR9 ligands but not STING ligands

R848 induced secretion of all the cytokines examined in almost all of the individuals studied (Figure 1c), whereas only IFN-α, MCP-1 and IL-8 were robustly induced in all subjects in response to ODN M362 (Supplementary Figure S1, Figure 1d). After cGAMP stimulation, we detected the same profile of cytokines as the one observed in R848 stimulated whole-blood assays, excepted for MCP-1 and IL-8 which were less efficiently induced (Figure 1c, e; Supplementary Figure S1 and S2). Altogether, our data show that these different agonist ligands induced biologic responses up to 1000-fold greater than the null conditions, for a selected set of cytokines, including IFN-α, which were reproducibly induced in most donors tested. Strikingly, sex differences were limited to the production of IFN-α induced in response to the TLR7/8 ligand R848 and the TLR9 ligand ODN M362 (Figure 1c, d; Supplementary Figure S3). Interestingly, the significant female preeminence for the production of IFN-α induced by the TLR9 ligand CpG-C ODN M362 has never been reported before. Robust production of IFN-α was also induced by the STING ligand cGAMP (Figure 1e). Although we noticed a trend toward an increased production in male, this was not significant. No sex-bias was found for any of the other analytes induced by cGAMP (Figure 1e).

We next investigated the effects of aging on interferogenesis in both sexes by subdividing individuals into four age-classes (from 19 to 39, from 40 to 59, from 60 to 79, over 80) (Supplementary Figure S4). IFN-α production, upon stimulation with R848 or ODN M362, was significantly higher in the two youngest groups <60, compared to the two oldest ones (60-80 and >80) (Supplementary Figure S4a, b). The blunted IFN-I responses to TLR-ligands in older ages contrasted with cGAMP-induced IFN-α production, which was significantly increased in the 60-80 subjects, compared to young 18-40 and middle-aged 40-60 subjects (Supplementary Figure S4c). Because the age of 60 appeared as a tipping point regarding IFN-I production in response to the TLR agonist ligands, we then assessed the effect of sex on IFN-I production below or above 60. Whereas the sex-bias in IFN-α secretion induced by R848 was robustly maintained for individuals under 60 and over 60, the female predominance preeminence in IFN-α production in response to ODN M362 was limited to the age group <60 (Figure 1f, g). We observed a significant reduction with aging in IFN-α secretion induced by the endosomal TLR ligands, R848 or ODN M362. This reduction was observed in both males and females (Figure 1f, g). By contrast, cGAMP-induced IFN-α production remained stable with aging in both sexes (Figure 1h).

Together, our data revealed a striking and selective influence of sex on IFN-α production driven by innate cell stimulation through TLR7 or TLR9, but not through the STING-IRF3 pathway. As already mentioned, no sex bias was observed regarding the production of IL-1ß, IL-6, TNF-α, IFN-γ, MCP-1 and IL-8, in response to the different types of ligands tested.

### The female preeminence in TLR7-driven IFN-α production is robustly maintained in elderly subjects

We first confirmed that pDCs were the main source of IFN-α in PBMCs stimulated with the TLR ligands in both sexes. As shown in Supplementary Figure S5 a-b, pDCs represented 0.5 % of total PBMCs in average. Selective depletion of pDCs before stimulation resulted in >10-fold reduction in pDC frequencies (Supplementary Figure S5b), which was associated with the inhibition by more than 95% of the production of IFN-α by PBMCs stimulated with R848 or ODN M362 (Supplementary Figure S5c, d). By comparing the effect of sex on the production of IFN-α in a sub-cohort with age > 80 (Figure 1A), we still observed higher IFN-α production in female blood cells stimulated with R848 compared to males (Student’s t test; p = 0.0019) (Figure 2a). No sex bias was observed in response to TLR9- or STING-stimulation (Figure 2b and c). Together, these results show that the superiority of female pDCs to produce IFN-I is robustly maintained in the elderly in response to TLR7 agonist ligands.

**Figure 2:**
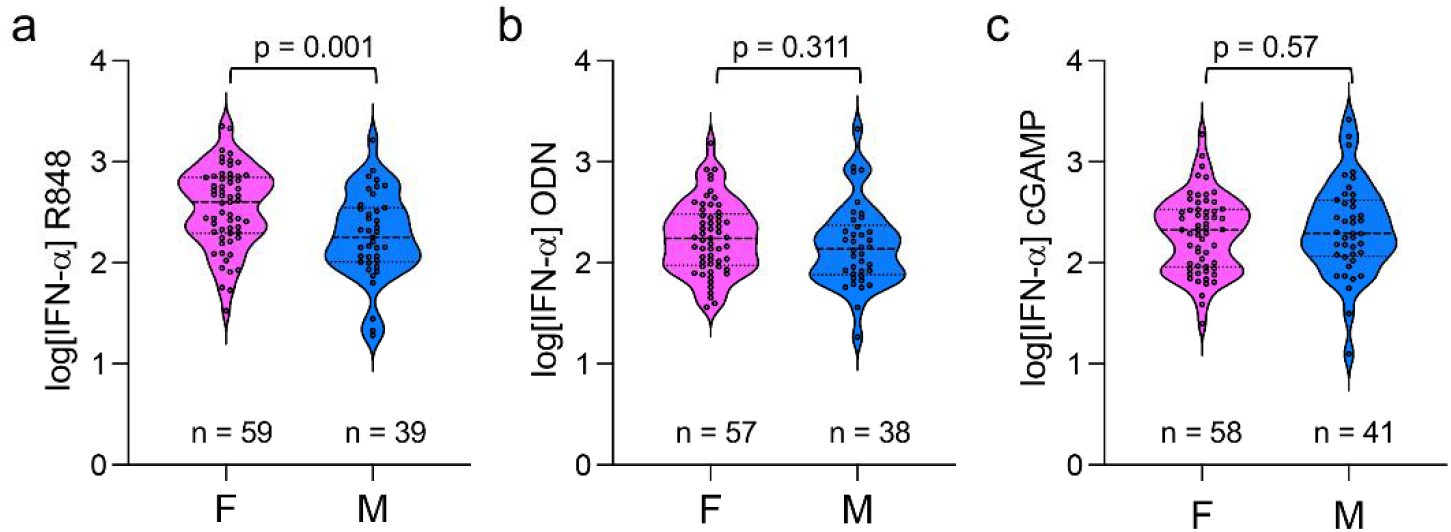
The female predominance in TLR7-driven production of IFN-α is robustly maintained in elderly subjects above 80. (a-c) From the source groups depicted in Fig. 1a (age range >80), the production of IFN-α is shown in response to R848 (a), ODN M362 (b) and cGAMP (c) by sex, female (F), male (M). Statistical differences between groups were analyzed using the unpaired *Student’s t*-*test*.

### Genetic polymorphisms in TLR7 and CXorf21 loci do not affect the sex-bias in type-1 interferon production

The *TLR7* polymorphism rs179008 (c.32A>T) has been recently identified as a sex-specific eQTL, associated with reduced expression of TLR7 protein and blunted IFN-α production by pDCs, selectively in females [21]. Another functional SNP in the 3’ UTR of *TLR7*, rs3853839, has been reported to be associated with an increased risk of developing SLE specifically in men [36]. Additionally, in European populations, polymorphism in the CXorf21 gene has been shown to be strongly associated with SLE in women, and SLE flares are associated with increased expression of CXorf21 mRNA in blood cells [37, 38]. We therefore decided to check the possible impact of these three X-linked frequent SNPs on the sex differences in the TLR7/9-induced IFN-α secretion. The allele frequencies of the *TLR7* SNP rs179008 rs3853839, and CXorf21 rs887369 were analyzed in our cohort (men n=135; women n=163). No significant bias in minor allele frequencies was observed between our cohort and the European 1000 genome project cohort (Supplementary Table S1), and the overall repartition of each allele between men and women was not significantly different from the calculated ones (Supplementary Table S2). We then selected among our cohort those subjects expressing the major alleles for each SNPs, and compared IFN-α production between men and women (Figure 3). For both *TLR7* rs179008 (AA vs A0) and rs3853839 (CC vs C0) major allele carriers, the female superiority in IFN-α production was maintained, not only in response to TLR7 ligands but also in response to TLR9 stimulation (Figure 3a, b). Similar results were observed comparing *CXorf21* rs887369 C/C women with C0 men, but the sex bias in IFN-α production was significantly maintained only for R848 stimulatory condition but not in response to TLR9 ligand M362 (Figure 3c). Of note, cGAMP-induced IFN-α production was significantly upregulated in C0 males compared to CC females, although the effect was modest and could result from a sampling effect (Figure 3c). Together, these results show that confounding factors such as the presence of frequent X-linked eQTL of *TLR7* and *CXOrf21/TASL* genes do not contribute to sex bias in the production of IFN-α by pDCs stimulated through either TLR7 or TLR9.

**Figure 3:**
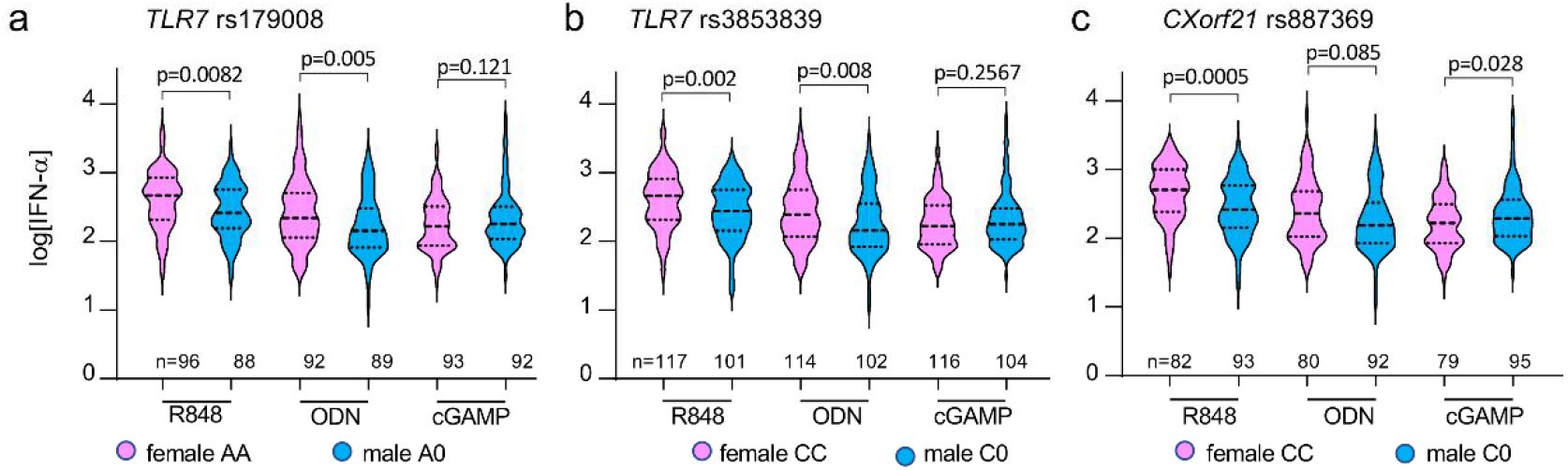
Impact of TLR7 and CXorf21 SNPs on the IFN-α response to the three PPR ligands studied. Individuals were genotyped for two TLR7 SNPs (rs179008 (Gln11Leu) and rs3853839 (in 3′ UTR) (a, b) and the rs887369 CXorf21 SNP (c). For each SNP, the IFN-α responses observed for female homozygous for the most frequent alleles were compared to the responses of males having the corresponding alleles. The results are given in log_10_ of the concentrations (pg/ml) with background (without ligands) subtracted. The unpaired *Student’s t*-*test* was used to compare observations between male and female groups.

### CMV status interferes with the age-related decrease in TLR7-driven IFN-I production but not with the sex bias

CMV is a confounding factor for the effect of age and we recently reported that the frequency of circulating pDCs significantly decreased with age in CMV-positive (CMV^pos^) but not in CMV-negative (CMV^neg^) individuals [34]. When we stratified our results according to the CMV serological status, we observed similar IFN-I response upon R-848 stimulation between CMV^pos^ (n= 115) and CMV^neg^ (n = 189) individuals (Figure 4a). As expected, the numbers of CMV^pos^ subjects increased with age in our cohort in both sexes (Supplementary Figure S6), which correlated with a reduction in TLR7-driven interferogenesis (p < 0.0001 and R = 0.279) (Figure 4b). By contrast, the correlation between age and response to R848 was not significant (p = 0.095 and R = 0.154) in CMV^neg^ individuals (Figure 4b). These results are consistent with the observation that CMV^pos^ status and age are associated with a decline in pDCs [34, 39], which represent the main source of IFN-α in response to R848 or CpG-C stimulation of PBMCs [20, 29, 30, 32], as shown in Supplementary Figure S5. Of note the sex-difference in R848-driven IFN-α production was still observed in CMV^neg^ or CMV^pos^ subjects (Figure 4c). Thus, although CMV seropositivity was associated with a blunted IFN-α response to R848 in both sexes, the female predominance in TLR7-driven production of IFN-α was robustly maintained.

**Figure 4:**
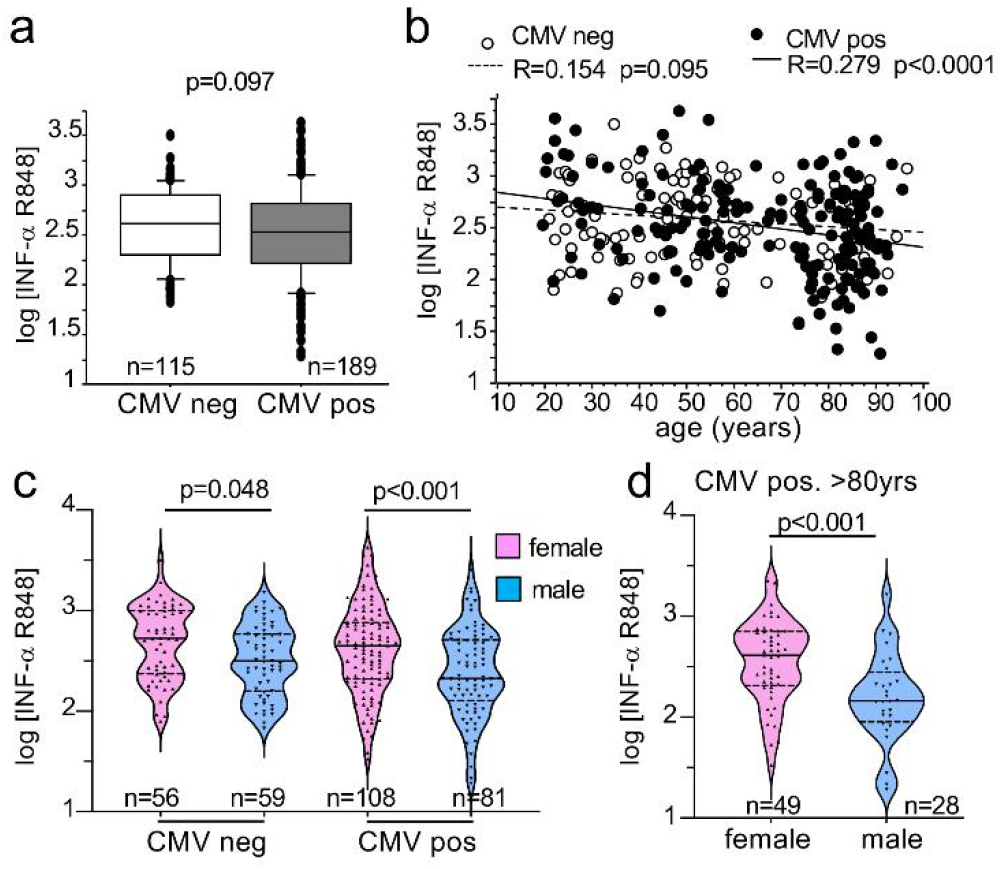
Influence of CMV serological status on the decrease of IFN-α release with aging. IFN-α release expressed as in Figure 1 in response to stimulation with R848 is shown according to the CMV status: CMV positive (CMV pos) and CMV negative (CMV neg) individuals (a) or as a function of age (b). The Pearson’s R coefficient and correlation probabilities were calculated by means of XLSTAT. Panel (c) shows the IFN-α production in CMV neg and CMV pos individuals by sex. (d) Production of IFN-α in CMV pos elderly subjects above 80. Differences between groups were analyzed using the Student’s t-test (a, d) or using a one-way ANOVA test followed by Sidak’s multiple comparisons test (c).

### Age-related changes in circulating pDCs and monocytes differ between sexes

We next investigated the effect of sex and aging on blood pDCs, monocytes and DCs numbers. We observed a significant reduction in pDCs numbers in subject >60 in females but not in males. Of note, in people <60, we observed a trend towards higher numbers of pDCs in females compared to males, although this was not significant (Figure 5a). In agreement with previous works, we also observed higher numbers of monocytes in male subjects >60 compared to age-matched females or males below 60 [40]. Because blood DC counts were not affected by age and sex (Figure 5a), we focused our correlation analysis on pDCs and monocytes. IFN-α productions in response to R848 or ODN M362 were highly correlated together and with the absolute number of blood pDCs (Student’s test: R = 0.474; p <0.0001 and R = 0.509; p <0.0001, respectively) (Figure 5b, c; Supplementary Figure 3). Because the IFN-α response to R848 or ODN M362 stimulations were correlated with the absolute number of pDC per mm^3^, we calculated the residual values (the difference between the observed IFN-α concentration and the predicted value from the linear regression of IFN-α response as a function of pDC number). The residual values were significantly higher in females than in males, particularly in subjects above 60 (Figure 5d, e). In response to TLR7 ligands, the residual values in women were above 0 in all classes of age (<60 and >60) and superior to the ones observed in men (Figure 5d). A similar trend was found in response to TLR9 although the sex differences was significant only for the age class >60 (Figure 5e). Together, these results indicate that the sex differences in pDC-derived IFN-α cannot be simply explained by sex differences in circulating pDC numbers, but rather reflect sex-specific intrinsic differences in the functional responses of pDCs in agreement with previous works [20, 21, 25, 41].

**Figure 5:**
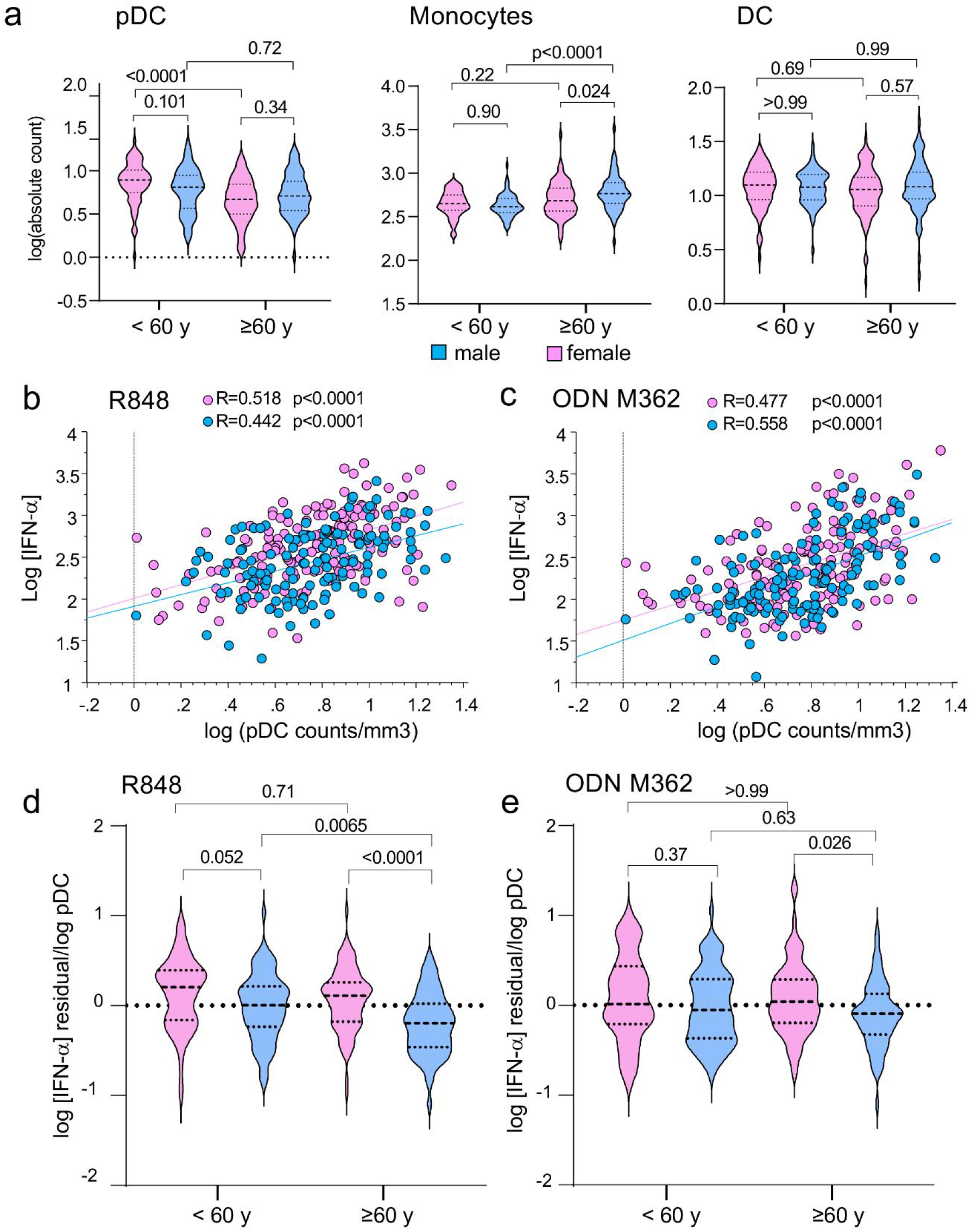
Aging-related changes in pDCs and monocytes differ between sexes. (a) Absolute cell counts of pDC, mDC and monocytes per mm^3^ were calculated from FCM data as described in materials and methods. Absolute counts per mm^3^ (given in log_10_) are compared in males and females for individuals under or above 60. (b, c) Correlation between log_10_ IFN-α release in response to R848 (b) or ODN M362 (c) stimulation and log_10_ absolute numbers per mm^3^ of plasmacytoid dendritic cells. Pearson’s correlation coefficients “ *R”* and probabilities of correlation were calculated by means of Statview software. (d, e) Residual values (difference between the observed IFN-α concentration and the predicted value) were calculated from the linear regression of IFN-α response as a function of pDC number shown in (b) and (c), respectively. Differences between groups were analyzed using a one-way ANOVA test followed by Sidak’s multiple comparisons test (a, d, e).

### CD14^+^ classical monocytes are the main source of STING-mediated IFN-α production in both sexes

As shown in Supplementary Figure S3, the IFN-α response to cGAMP did not correlate with the IFN-α responses to R848 or ODN M362, suggesting that IFN-α is produced by distinct cell populations in both systems. Indeed, although, pDCs express cGAS/STING and can respond to cGAMP by producing IFN-α [42], in our dataset, the IFN-α response to cGAMP correlated with the number of monocytes rather than with the number of pDCs (Supplementary Figure S3 and Figure 6b). As expected from previous work, ageing was correlated with a steadily increase of monocytes, in both sexes but particularly in men (Figure 5a and Figure 6a). As cGAMP stimulated IFN-α secretion was positively correlated with the numbers of monocytes (Figure 6b), we determined the monocyte contribution to the production of IFN-α in cGAMP-stimulated PBMCs. As shown in Figure 6c, CD14-depleted PBMCs exhibited a marked reduction in IFN-α production in response to cGAMP compared to mock-depleted PBMCs. The efficiency of CD14^+^ monocyte depletion was above 98% (Supplementary Figure S7). As expected, IFN-α production in R848 stimulated PBMCs was not affected by CD14 cell depletion, as in these conditions, IFN-α was mainly produced by pDCs (Supplementary Figure S5). Indeed, IFN-α or IP-10 (CXCL10) were readily detectable in monocyte-enriched cultures in response to cGAMP stimulation than PBMCs (Figure 6d, e). By calculating the ratio of the median actual values of cytokine concentrations, we estimated that monocytes-enriched cultures produced 4- to 5-times more IFN-α or IP-10 (CXCL10) in response to cGAMP stimulation as compared to PBMCs on a per-cell basis (Figure 6d, e). cGAMP-induced IFN-α or IP-10 production were not significantly different between men and women. Lastly, purified monocytes were assessed for intracellular expression of IFN-α upon cGAMP stimulation. We could directly visualize IFN-α production in a small but clearly distinct population of cGAMP-stimulated monocytes (comprising 4% to 7% of monocytes) (Figure 6f, Supplementary Figure S8). Again, no sex differences were observed in the frequency of IFN-α^+^ monocytes in response to cGAMP (Figure 6g). Together, these results show that monocytes represent the main cellular source of IFN-α in cGAMP-stimulated PBMC cultures from both men and women. Moreover, we found no evidence for a sex-biased response in the STING-signaling pathway in human monocytes.

**Figure 6:**
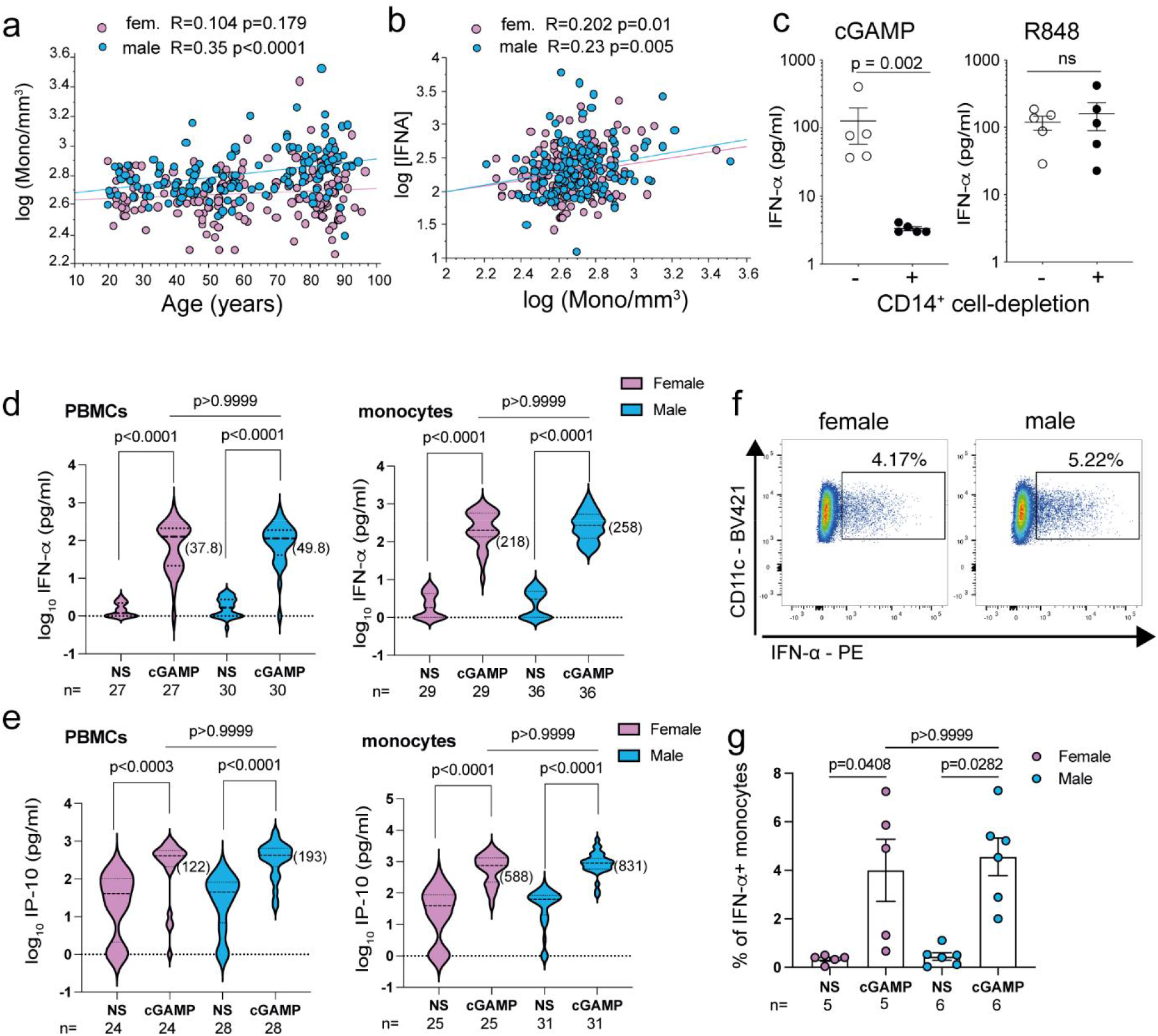
Monocytes are the main source of IFN-α in response to cGAMP. Impact of aging on monocyte counts (a) and IFN-α response (b) after cGAMP stimulation for males and females. Correlation coefficients “ R” and probabilities of correlation were calculated using Pearson test. (a) Correlation between the absolute number of monocytes/mm^3^ (log_10_) and age (years). (b) Correlation between the cGAMP-stimulated release of IFN-α (log_10_) and the absolute number of monocytes/mm^3^ (log_10_). (c) PBMC from 5 donors were depleted of CD14^+^ cells (> 98% depletion) using magnetic particles or mock-depleted and stimulated with cGAMP (20 µg/ml) or R848 (1 µg/ml) for 24 hours. IFN-α production was measured by ELISA. Paired Student’s *t*-test was used for statistical analysis. (d, e) Total PBMCs (5 × 10^5^ cells/well) or monocyte-enriched cells (% CD14^+^ cells > 77% at 3 × 10^5^ cells/well) were stimulated with cGAMP or left untreated (NS). The production of IFN-α (d) or IP-10/CXCL-10 (e) was then assessed in 24 hour-supernatants. Median values in pg/ml for [IFN-α] or [IP-10] concentrations normalized to 3 × 10^5^ cells/well are shown in parentheses. (f, g) Intracellular analysis of IFN-α synthesis in 18 hour-stimulated monocytes, gated as CD11c^pos^ HLA-DR^pos^ cells, from male or female donors. Brefeldin A was added for the last 3 hours of culture. Frequencies of IFN-α^pos^ monocytes are shown from individual female or male donors stimulated or not with cGAMP. Statistical differences between groups were assessed using the Kruskal-Wallis’s test corrected for multiple comparisons.

## Discussion

In this study, we investigated the impact of sex and aging on innate cytokine release in a whole-blood assay comprising 145 men and 165 women aged 19 to 97 screened to rule out confounding diseases and treatments. IFN-α was the only cytokine exhibiting sex differences among the 7 analytes investigated. We showed that the female preeminence in TLR7-induced IFN-α production was strongly maintained over the ages, including in subjects over 80, and persisted even after accounting for confounding factors such as CMV seropositivity or polymorphisms of two X-linked genes respectively encoding TLR7 and the TASL/CXorf21 adapter molecule. Interestingly, we also provided the first evidence for a female’s sex-biased response in IFN-α production induced by TLR9, not only in younger subjects <60, but also in subjects >60 after correcting for circulating pDC numbers. This enhanced IFN-I production observed in female PBMCs stimulated through TLR9 was missed in previous studies [18], probably due to the use of a less powerful TLR9 agonist ligand unlike the one we used in the present work [35]. On the contrary, cGAMP/STING-induced interferogenesis did not differ significantly as a function of sex. The cGAMP/STING-induced IFN-α production neither correlated with the IFN-α response induced by the TLR7/8/9 ligands nor with the number of circulating pDCs. Instead, we found strong positive correlation with the number of monocytes, and we provided evidence that monocytes were the major source of IFN-α upon cGAMP activation in both sexes. This may explain the trend towards enhanced STING-induced interferogenesis with aging, particularly in males, which could result from the age-related changes in circulating monocytes, rather than sex- or age-specific functional differences in monocyte biology.

Sex bias in immune cell proportions exists in healthy adult human subjects [43] as well as elderly people [34, 40, 44]. Robust differences in the blood transcriptome of healthy men and women have been recently described across worldwide human populations [43, 45, 46]. Such sex-specific expression signatures have been described identifying robust changes in immune cell proportions with aging, and predicted antibody response to influenza infection in a sex-specific manner [43]. In our study, we observed a significant increase in monocyte numbers in older subjects (60+), which was more prominent in older males compared to older females. Such age-related changes have been observed by others and found to be associated with up-regulation of inflammatory markers of innate immunity such as IL-1RA and IL-6 [45, 46]. Of note, we observed a trend towards higher production of IFN-α after cGAMP stimulation particularly in older ages (60+), which was correlated with a raise in monocyte numbers with aging. Although it has been shown that both pDCs and monocytes expressed high levels of the cGAS and STING proteins as compared to B and T cells, the cellular origin of IFN-α from cGAMP-stimulated PBMCs has never been characterized [42]. Herein, we provide the first evidence that monocytes represent the major source of IFN-α upon stimulation of the STING-pathway in whole PBMCs. We also show by intracellular staining that cGAMP stimulates IFN-α in a discrete but sizable population of monocytes, suggesting that some functional heterogeneity may exist in human monocytes regarding STING-driven IFN-I production. Contrary to TLR-agonist ligands, no major sex bias was found when examining IFN-I production induced by cGAMP, suggesting minor cell-intrinsic differences in STING activation between monocytes from men and women.

Blood pDCs are the exclusive source of IFN-α upon whole PBMC stimulation with either TLR7- or TLR9-specific ligands [20, 29, 30, 32]. In agreement with previous works [39], we also observed a marked reduction in the numbers of circulating pDCs in elderly subjects. A robust feature which has been found associated with blunted type I IFN production in response to influenza infection [47, 48]. This negative impact of aging on circulating pDCs was much more pronounced in females compared to males. Whether this is due to reduction in estrogen levels in female will deserve further investigations. Despite this reduction in blood pDCs, sex-differences in IFN-α production still persisted in response to TLR7, and also in response to TLR9 when adjusted to the numbers of circulating pDCs. Thus, the female bias in the production of IFN-α cannot be simply explained by the change in pDCs numbers and likely stems from cell-intrinsic differences due to estrogen-signaling [20, 41, 49] and/or X chromosome complement effects [25, 50], or both [41]. In favor of a genetically driven mechanism controlling pDC-derived IFN-I production, it has been recently reported that *TLR7* escapes from X chromosome inactivation in female immune cells [9, 50], and that all IFN-α subtypes as well as IFN-ß were transcribed at higher levels in female pDCs with biallelic expression of *TLR7*, compared to monoallelic cells [25]. These genetic effects associated with X chromosome dosage are likely to persist after menopause in aged women, and could contribute to the female preeminence in the TLR7-driven production of IFN-α that is robustly maintained through ages, including over 80 years old. Furthermore, the higher expression of mRNA for IFN-I in pDCs with biallelic expression of *TLR7* [25] suggest that such pDCs would belong to a group of cells prone for initiation of IFN-I secretion not only in response to TLR7 ligands but also TLR9 stimulation [51]. This could explain why we still observed a female superiority in TLR9-driven IFN-α production at older ages (60+), when correcting for circulating pDC numbers.

Our study presents some limitations. We used different PRR agonist ligands for which the kinetics in IFN-α production are likely different. However, kinetics issue is unlikely to influence the sex differences in IFN-α production driven by TLR7 ligands, as we recently reported that the sex bias in IFN-α production by PBMCs stimulated with resiquimod (R-848) or with imiquimod (R-837) was readily observed at 6-, 12-, and 24-hours post-stimulation [21]. This is also unlikely to explain the lack of sex bias in IFN-α production at 24 hours in response to cGAMP, as similar results were obtained between male and female by measuring the frequency of IFN-α-producing monocytes by intracellular staining at earlier time point. We cannot generalize our observation to IFN-β or type III IFN subtypes, as our assay captured mostly IFN-α subtypes, including IFN-α1/13, 2,4−8, 10, 14, 16, and 17. Finally, our results were obtained in participants which were selected to equilibrate both sexes in each 20-year age strata, with a particular emphasis in the recruitment of similar numbers of male and female at the age >80. Thus, regarding the repartition by sexes our cohort is not representative of the general population.

Taken together, our data demonstrate that the female predominance of blood pDCs to produce IFN-I in response to TLR7 agonist ligand is maintained in elderly women. This is supportive of the general hypothesis that this pathway could represent an important contributor of the enhanced protection of females against different class of viruses including SARS-CoV2 [5, 6]. In contrast, STING-induced IFN-I production was not sex-biased. Interestingly, we show here that monocytes were the main source of IFN-α in cGAS-stimulated PBMCs. Thus, targeting the STING pathway in monocytes could represent a promising strategy to improve immune surveillance and control of tumors [52], or to optimize vaccine design [53-55] with similar efficacy in both sexes. Finally, it is becoming increasingly evident that the cGAS-cGAMP-STING pathway connects persistent DNA damage to autoinflammation and senescence [56, 57]. Whereas the IFN-I production by the cGAS-STING pathway in response to viral and bacterial DNA is generally protective by mediating immunosurveillance, excessive and chronic stimulation of this pathway can also be deleterious [58]. Because myeloid cells such as monocytes accumulate with aging in both sexes, but more specifically in males, investigating the contribution of STING activation in monocytes in sex-differences in inflammaging [45] will warrant further investigation.

## Data Availability

All data produced in the present study are available upon reasonable request to the authors

## Contributors

AB, NC and JD, conceived and designed the clinical trial, performed experiment and analyzed the data. JCG and AB conceptualized the present study, analyzed the data and supervised experiments. BPL, KG and PAA contributed by FMC analysis of immune cells in blood samples. CC, FA, SL performed SNP analysis and experiments to analyze the origin of IFN-I producing cells, and interpreted the data. AB and JCG wrote the manuscript with input from their co-authors. All authors read and approved the final version of the manuscript. AB and JCG have accessed and verified the data and were responsible for the decision to submit the manuscript.

## Data Sharing

All of the data generated or analyzed during this study are included in this article and the Supplementary informations. All data supporting the findings of this study are also available upon reasonable request from the corresponding authors [A.B. or J-C.G.].

## Acknowledgments

We acknowledge the anonymous subjects who participated to this study for their generous blood donation. This work was supported by grants from Région Midi-Pyrénées, France (#12052910), University Paul Sabatier (UPS Toulouse 3), and by the Inspire Program from the Région Occitanie/Pyrénées-Méditerranée (#1901175) and the European Regional Development Fund (MP0022856).

## Supplementary materials

Supplementary material is associated with this article.

