## Supplementary figures and tables for "Monocytes are the main source of STING-mediated IFN-α production": CLEAN COPY_Supplemental Information.pdf

### Contents

| | Page |
|---|---|
| **Supplementary Figure S1**: Raw cytokine responses in the presence or absence of PRR ligands | 2 |
| **Supplementary Figure S2:** Frequencies of non-responding individuals by sex | 3 |
| **Supplementary Figure S3.** Correlation matrix of investigated read-outs | 4 |
| **Supplementary Figure S4.** Age-related changes in IFN-α production | 5 |
| **Supplementary Figure S5.** pDC depletion assay | 6 |
| **Supplementary Figure S6.** Age-related changes in CMV serostatus by sex | 7 |
| **Supplementary Figure S7.** Monocyte depletion assay | 7 |
| **Supplementary Figure S8.** Flow cytometric analysis of IFN-α-producing monocytes | 8 |
| **Supplementary Table S1:** Allelic frequency of rs178008, rs3853835 and rs887369 in comparison to the European 1000 Genome project cohort | 9 |
| **Supplementary Table S2.** Repartition of rs178008, rs3853835 and rs887369 genotypes by sex | 10 |

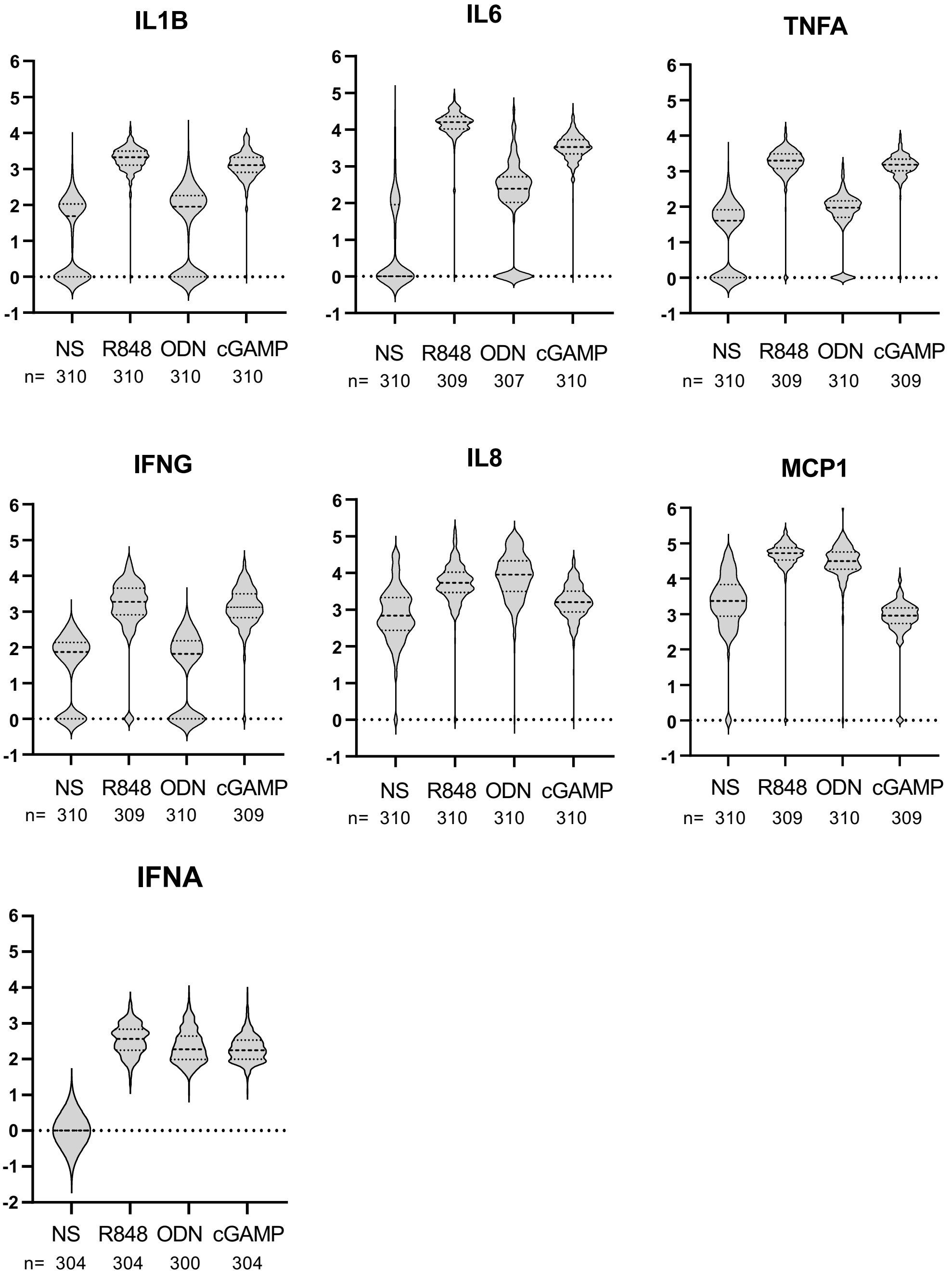


**Supplementary Figure S1. Raw cytokine responses in the presence or absence of PRR ligands**. The seven cytokine concentrations in plasma were measured after 24 h of stimulation. A control tube without any stimulus was processed in parallel with the tubes containing the three stimuli. For interferon alpha the measurement was carried out by a sandwich ELISA test (Mabtech), for the other cytokines the measurements were carried out by HTRF (Biosys). The results are given as a $log_{10}$ of the concentration measured after stimulation (in pg/mL). When the concentration was under the limit of detection, the value in $log_{10}$ is arbitrarily represented as null. When the concentration was above the upper limit of quantification, the value in $log_{10}$ is arbitrarily represented as 6.

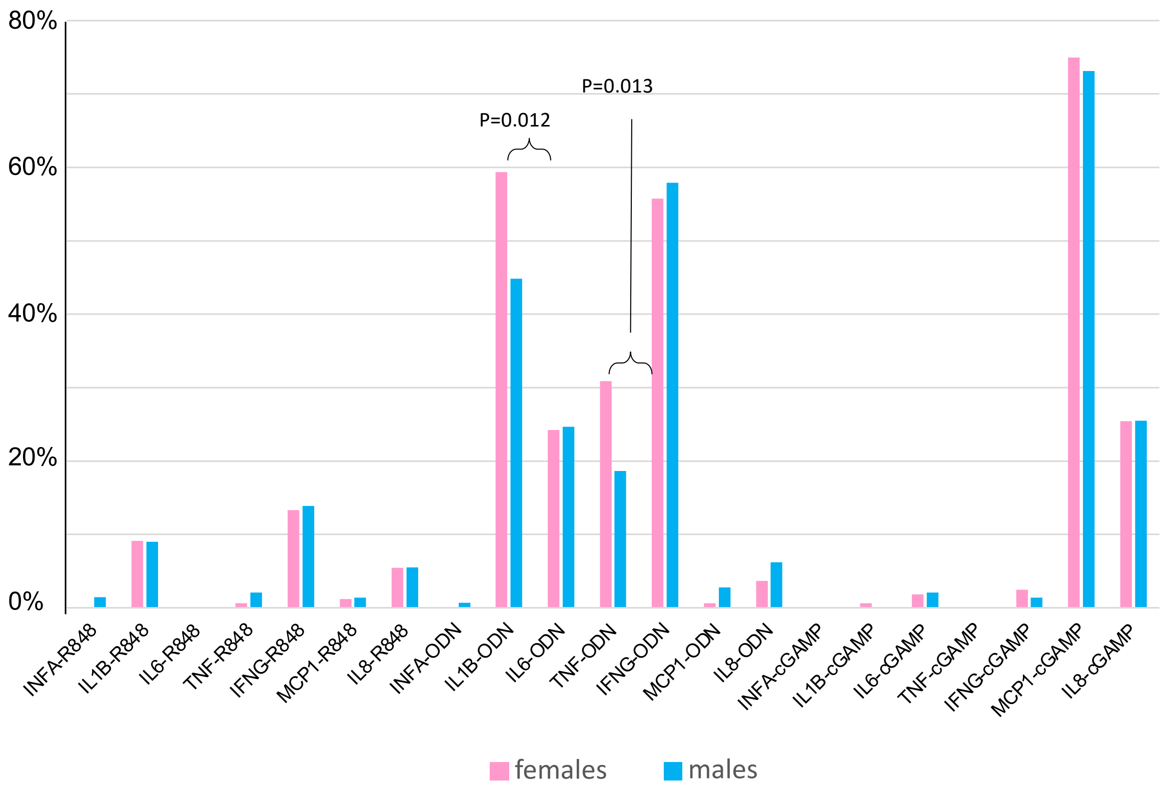


**Supplementary Figure S2. Frequencies of non-responding individuals by sex.** For each stimulus and each cytokine, the histograms give the proportion of non-responding males and females. Significant differences between males and females are shown (exact probabilities are calculated by means of Fisher's exact test.

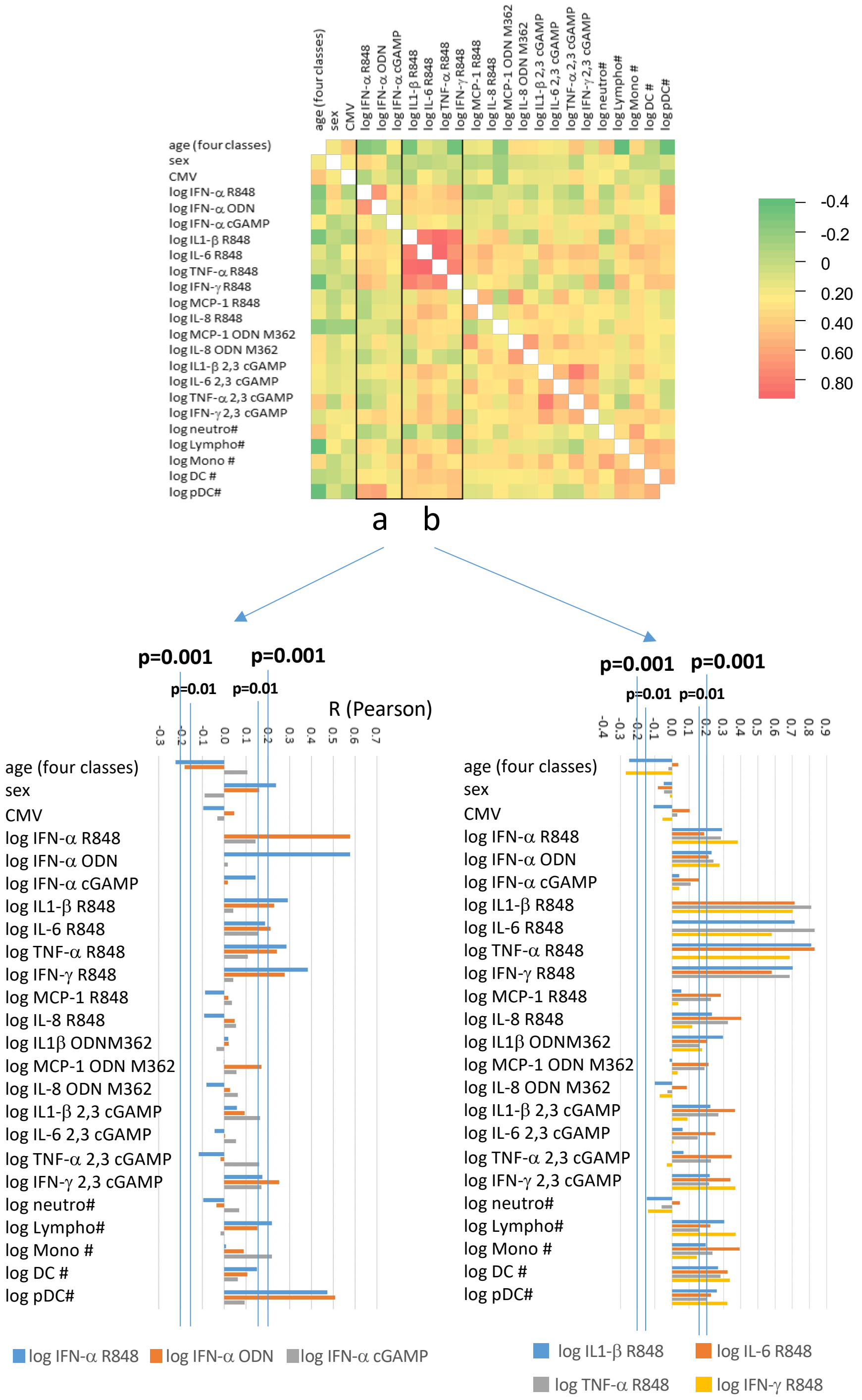


**Supplementary Figure S3. Correlation matrix of investigated read-outs.** Pearson's correlation coefficients "r" and probabilities of correlation were calculated by means of XLSAT software. Individuals were distributed in four groups of age (19 to 40, 40 to 60, 60 to 80, more than 80). The stimulated cytokine release for which high proportions of non-responding individuals were observed are not considered. Histograms "a" gives the "r" correlation coefficients between all parameters analysed and the IFN-α release signals observed after R848, ODN and cGAMP stimulation. Histogram "b" gives the "r" correlation coefficients between all parameters analysed and the IL1-ß, IL-6, TNF-α and IFN-γ release signals observed after R848 stimulation The probabilities were corrected by using the False Discovery Rate (FDR) as described by Benjamini-Hochberg. For histograms "a" and "b", the limits of significance of "r" at p=0.01 and p=0.001 are shown

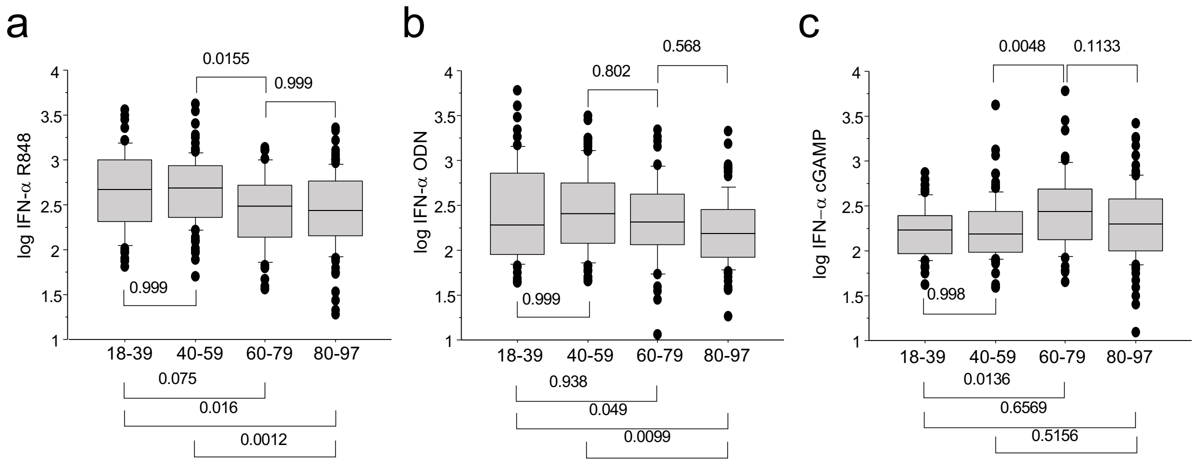


**Supplementary Figure S4. Age-related changes in IFN-α production in response to PRR ligands.** Results were obtained as described in Supplementary Figure S1 and expressed as log10 of the concentration (pg/ml) measured after stimulation with R848 (a) ODN M362 (b) or cGAMP (c). Groups were compared by a one-way ANOVA test followed by Sidak's multiple comparisons test.

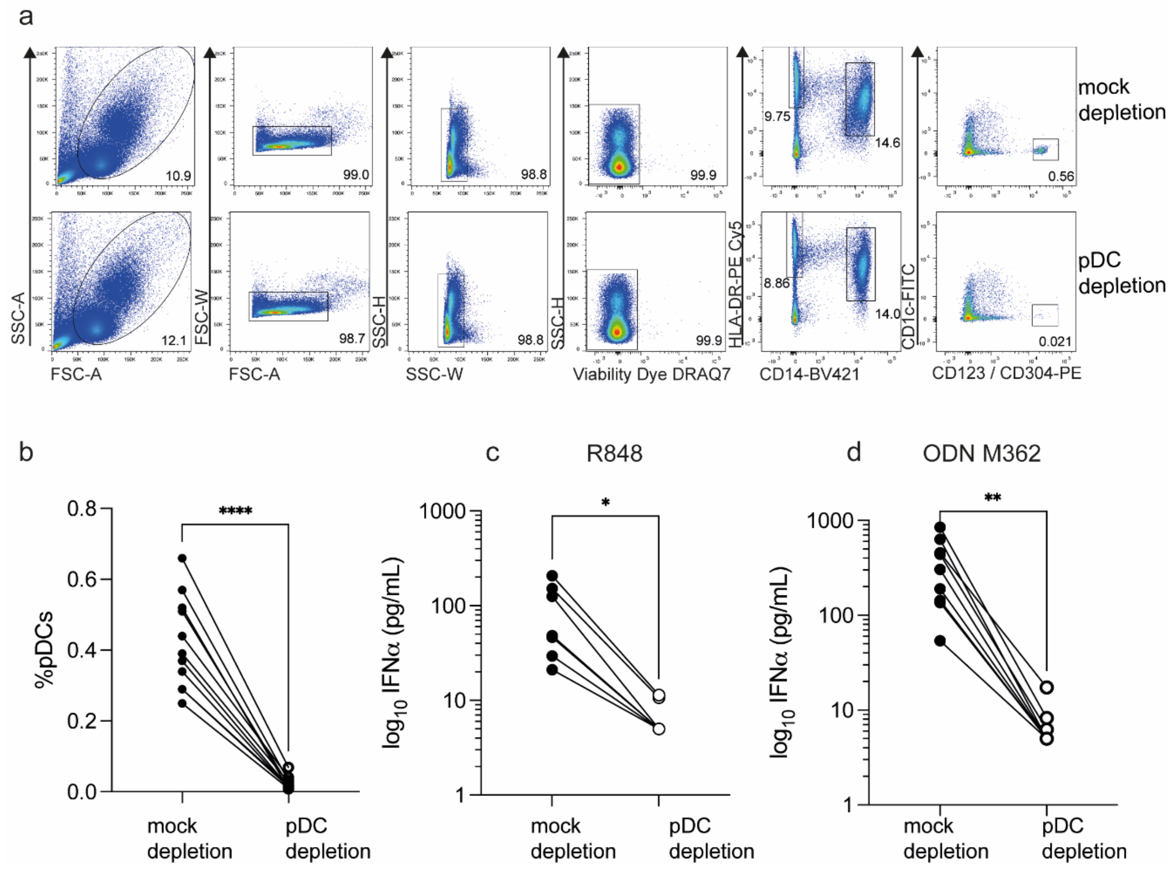


**Supplementary Figure S5. pDC-depletion assay.** Freshly isolated PBMCs from male and female donors were stained with BDCA4-PE and CD123-PE antibodies and then depleted using anti-PE microbeads (Miltenyi). (a) Gating strategy for pDC analysis. (b-d) PBMCs were depleted of BDCA4$^{+}$ CD123$^{+}$ cells using magnetic particles or mock-depleted and stimulated with 1 μM R848 (c), 1 μM ODN M362 (d) for 24 hours. IFN-α production was measured by ELISA. Statistical differences between groups were assessed using the paired Wilcoxon signed-rank test. The efficiency of pDC-depletion was > 95% in average.

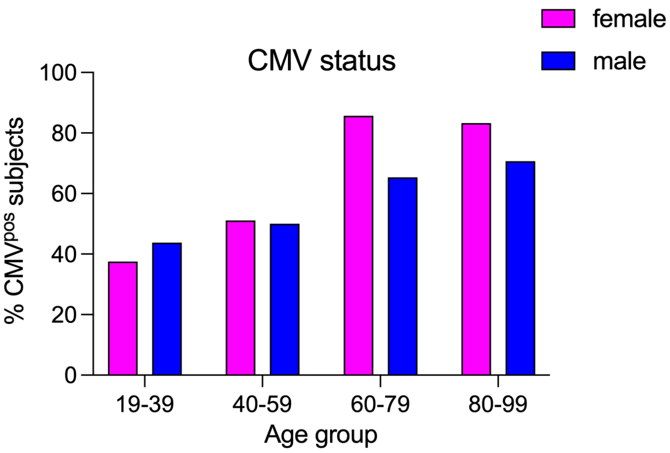


**Supplementary Figure S6. Age-related changes in CMV serostatus by sex**. For each age groups presented in Figure 1a the % of CMV positive subjects is shown by sex.

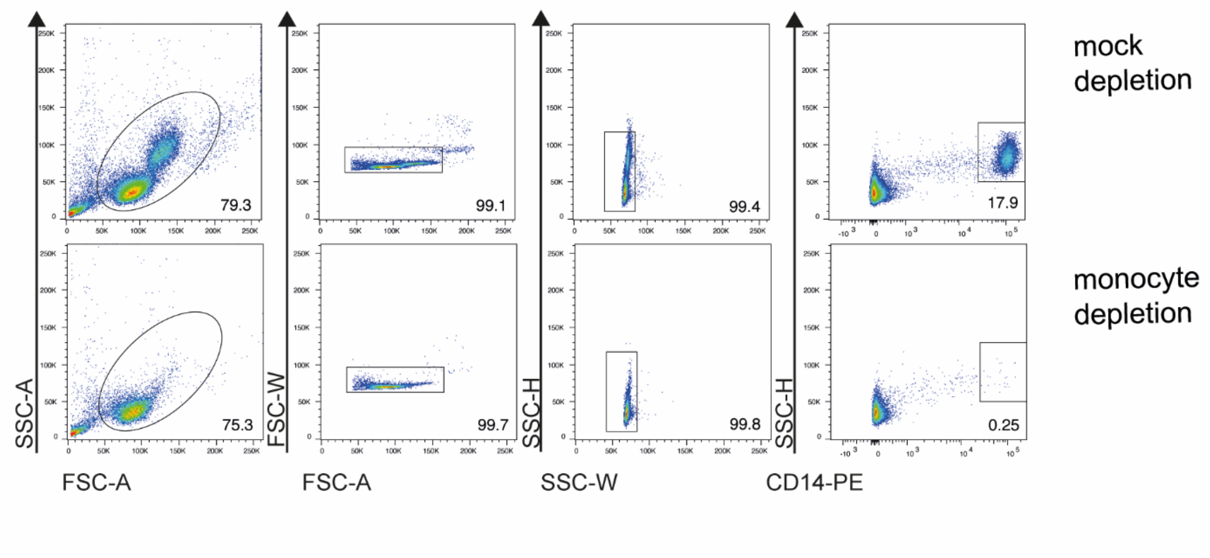


**Supplementary Figure S7. Monocyte depletion assay**. Freshly isolated PBMCs were depleted of $CD14^{+}$ cells using magnetic particles followed by autoMACS Pro running program Depletes, or mock depleted. The efficiency of $CD14^{+}$ cell depletion was more than 98%.

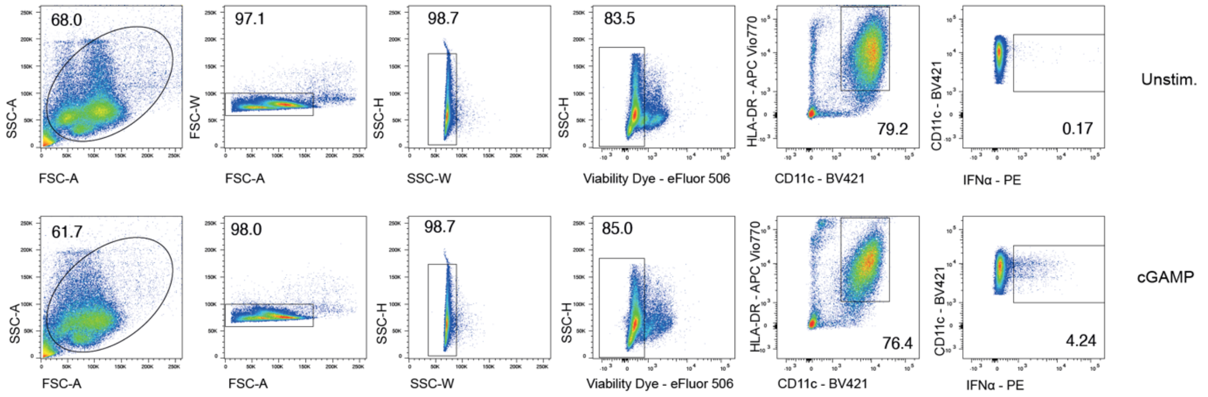


**Supplementary Figure S8. Flow cytometric analysis of IFN-α-producing monocytes.** Monocytes were purified from freshly isolated PBMCs using the EasySep$^{TM}$ human monocyte enrichment kit (StemCell technologies). Monocytes (purity > 77% CD11c$^{+}$DR$^{+}$CD14$^{+}$ cells) were stimulated for 18 hours with cGAMP 20 µg/ml in R10 complete medium, in the presence of brefeldin A for the final 3 hours. The cells were then harvested, surface stained, fixed, permeabilized, and intracellularly stained for IFN-α. Gating strategy to analyse the population of cytokine-producing human in HLA-DR$^{+}$ CD11c$^{+}$ monocytes in non-stimulated (upper panels) or cGAMP-stimulated (lower panels) cells is shown.

**Supplementary Table S1 :** Allelic frequency of rs178008. rs3853835 and rs887369 in comparison to the European 1000 Genome project cohort

| | SNP rs179008 (TLR7 exon 3) | |
|---|---|---|
| | A | T |
| 1000 genome project : all European | 0.77 | 0.23 |
| present study | 0.77 | 0.23 |

| | SNP rs3853839 (TLR7 3'UTR) | |
|---|---|---|
| | C | G |
| 1000 genome project : all European | 0.83 | 0.17 |
| present study | 0.84 | 0.16 |

| | SNP rs887369 (CXOrf21) | |
|---|---|---|
| | C | A |
| 1000 genome project : all European | 0.76 | 0.24 |
| present study | 0.73 | 0.26 |

**Supplementary Table S2:** Repartition of *TLR7* and *CXOrf21* SNP alleles among men and women.

| SNP rs179008 (TLR7 exon 3) | | | | | |
|---|---|---|---|---|---|
| | A/A | A/T | T/T | A/0 | T/0 |
| calculated | 97 | 58 | 8 | 104 | 31 |
| observed | 102 | 53 | 8 | 98 | 37 |
| Exact p: | | 0.82 | | 0.48 | |

| SNP rs179008 (TLR7 exon 3) | | | | | |
|---|---|---|---|---|---|
| | C/C | C/G | G/G | C/0 | G/0 |
| calculated | 116 | 43 | 4 | 114 | 21 |
| observed | 121 | 37 | 5 | 109 | 26 |
| Exact p: | | 0.76 | | 0,52 | |

| SNP rs887369 (CXOrf21) | | | | | |
|---|---|---|---|---|---|
| | C/C | C/A | A/A | C/0 | A/0 |
| calculated | 89 | 63 | 11 | 100 | 35 |
| observed | 86 | 68 | 9 | 100 | 35 |
| Exact p : | | 0.79 | | 1 | |

Legend of supplementary table 2: Exact probabilities were calculated by using the Fisher's exact test (males) or the Freeman-Halton extension of Fisher's exact test to calculate the (two-tailed) probability of obtaining a distribution of values in a 2x3 contingency table (females) (Soper 2021).

Reference: Soper, D.S. (2021). Fisher's Exact Test Calculator for a 2x3 Contingency Table [Software]. Available from https://www.danielsoper.com/statcalc.
